# Detection of SARS-CoV-2 infection by rapid antigen test in comparison with RT-PCR in a public setting

**DOI:** 10.1101/2021.01.22.21250042

**Authors:** Kathrine Kronberg Jakobsen, Jakob Schmidt Jensen, Tobias Todsen, Freddy Lippert, Cyril Jean-Marie Martel, Mads Klokker, Christian von Buchwald

## Abstract

**Background:** Rapid and accurate detection of SARS-CoV-2 infection is essential in limiting the spread of infection during the ongoing COVID-19 pandemic. The aim of this study was to determine the accuracy of the STANDARD Q COVID-19 Ag test (SD BIOSENSOR) by comparison with RT-PCR in a public setting.

**Method:** Individuals aged 18 years or older who had booked an appointment for a RT-PCR test on December 26-31, 2020 at a public test center in Copenhagen, Denmark, were invited to participate. An oropharyngeal swab was collected for RT-PCR analysis, immediately followed by a nasopharyngeal swab examined by the STANDARD Q COVID-19 Ag test (SD BIOSENSOR). Sensitivity, specificity, positive and negative predictive values of the antigen test were calculated with test results from RT-PCR as reference.

**Results:** Overall, 4697 individuals were included (female n=2456, 53.3%; mean age: 44.7 years, SD: 16.9 years); 196 individuals were tested twice or more. Among 4811 paired conclusive test results from the RT-PCR and antigen tests, 221 (4.6%) RT-PCR tests were positive. The overall sensitivity and specificity of the antigen test were 69.7% and 99.5%, the positive and negative predictive values were 87.0% and 98.5%. Ct values were significantly higher among individuals with false negative antigen tests compared to true positives.

**Conclusion:** The sensitivity, specificity, and predictive values found indicate that the STANDARD Q COVID-19 Ag is a good supplement to RT-PCR testing.

## Introduction

Rapid and accurate detection of SARS-CoV-2 infection is essential in limiting the spread of infection during the ongoing COVID-19 pandemic. The cornerstone of SARS-CoV-2 testing is real-time reverse transcriptase-polymerase chain reaction (RT-PCR) of an upper-respiratory specimen.

RT-PCR testing relies on centralized laboratory capacity and complex logistics, thus causing delays and bottlenecks with high sample numbers. Rapid antigen tests (RAT) can be performed onsite, are easy to administer, and the results are available within minutes. This enables an increased pace of testing and faster tracing of infected individuals. However, the accuracy of the RATs is questioned. The aim of this study was to determine the accuracy of the WHO EUL-approved STANDARD Q COVID-19 Ag test (SD BIOSENSOR) by comparison with RT-PCR in a public setting.

## Methods

Individuals aged 18 years or older who had booked an appointment for a RT-PCR test on December 26-31, 2020 at a public test center in Copenhagen, Denmark, were invited to participate. An oropharyngeal swab was collected for RT-PCR analysis, immediately followed by a nasopharyngeal swab examined by the STANDARD Q COVID-19 Ag test (SD BIOSENSOR). The criteria for positive RT-PCR test result were cycle threshold (Ct) ≤38 and ≥10. Participants were asked to fill out an online questionnaire regarding symptoms.

Sensitivity, specificity, positive and negative predictive values of the RAT were calculated with test results from RT-PCR as reference. A boxplot depicting difference in Ct values between participants with true positive and false negative RATs, including analysis for statistical difference by Wilcoxon test, were performed in R statistics (version 3.6.1).

## Results

Overall, 4697 individuals were included (female n=2456, 53.3%; mean age: 44.7 years, SD: 16.9 years); 196 individuals were tested twice or more.

Paired conclusive test results from the RT-PCR tests and RAT were accessible for 4811 tests, of these 221 (4.6%) RT-PCR tests were positive; 97 RT-PCR tests were missing or inconclusive (i.e. Ct>38). The overall sensitivity and specificity of the RAT were 69.7% and 99.5%, the positive and negative predictive values were 87.0% and 98.5% (Table 1). Changing the criteria of positive RT-PCR to Ct≤30 increased the sensitivity of the RAT to 81.1%.

**Table 1:** Agreement between RT-PCR test results and antigen test results overall, and for participants with and without symptoms.

| Overall |  |  |  |
| --- | --- | --- | --- |
|  | RT-PCR positive | RT-PCR negative | Total (%) |
| Antigen test positive (%) | 154 (3.2) | 23 (0.5) | 177 (3.7) |
| Antigen test negative (%) | 67 (1.4) | 4567 (94.9) | 4634 (96.3) |
| Total (%) | 221 (4.6) | 4590 (95.4) | 4811 (100) |
| Sensitivity | Specificity | Positive predictive value | Negative predictive value |
| 69.7% | 99.5% | 87.0% | 98.6% |
| With symptoms (self-reported) |  |  |  |
|  | RT-PCR positive | RT-PCR negative | Total (%) |
| Antigen test positive (%) | 67 (9.5) | 7 (1.0) | 74 (10.5) |
| Antigen test negative (%) | 18 (2.6) | 613 (87.3) | 631 (89.5) |
| Total (%) | 85 (12.1) | 620 (87.9) | 705* (100) |
| Sensitivity | Specificity | Positive predictive value | Negative predictive value |
| 78.8% | 98.9% | 90.5% | 97.1% |
| Without symptoms (self-reported) |  |  |  |
|  | RT-PCR positive | RT-PCR negative | Total (%) |
| Antigen test positive (%) | 29 (1.0) | 11 (0.4) | 40 (1.3) |
| Antigen test negative (%) | 30 (1.0) | 2938 (96.8) | 2968 (98.7) |
| Total (%) | 59 (2.0) | 2949 (98.0) | 3008* (100) |
| Sensitivity | Specificity | Positive predictive value | Negative predictive value |
| 49.2% | 99.6% | 72.5% | 99.0% |

\*Not all participants responded to the online questionnaire regarding symptoms

Among participants with self-reported symptoms and paired conclusive test results (n=705) the sensitivity and specificity of the RAT were 78.8% and 98.8%, and for participants without symptoms (n=3008) this was 49.2% and 99.6% (Table 1). Ct values were significantly higher among individuals with false negative RATs compared to true positives (Figure 1).

**Figure 1:**
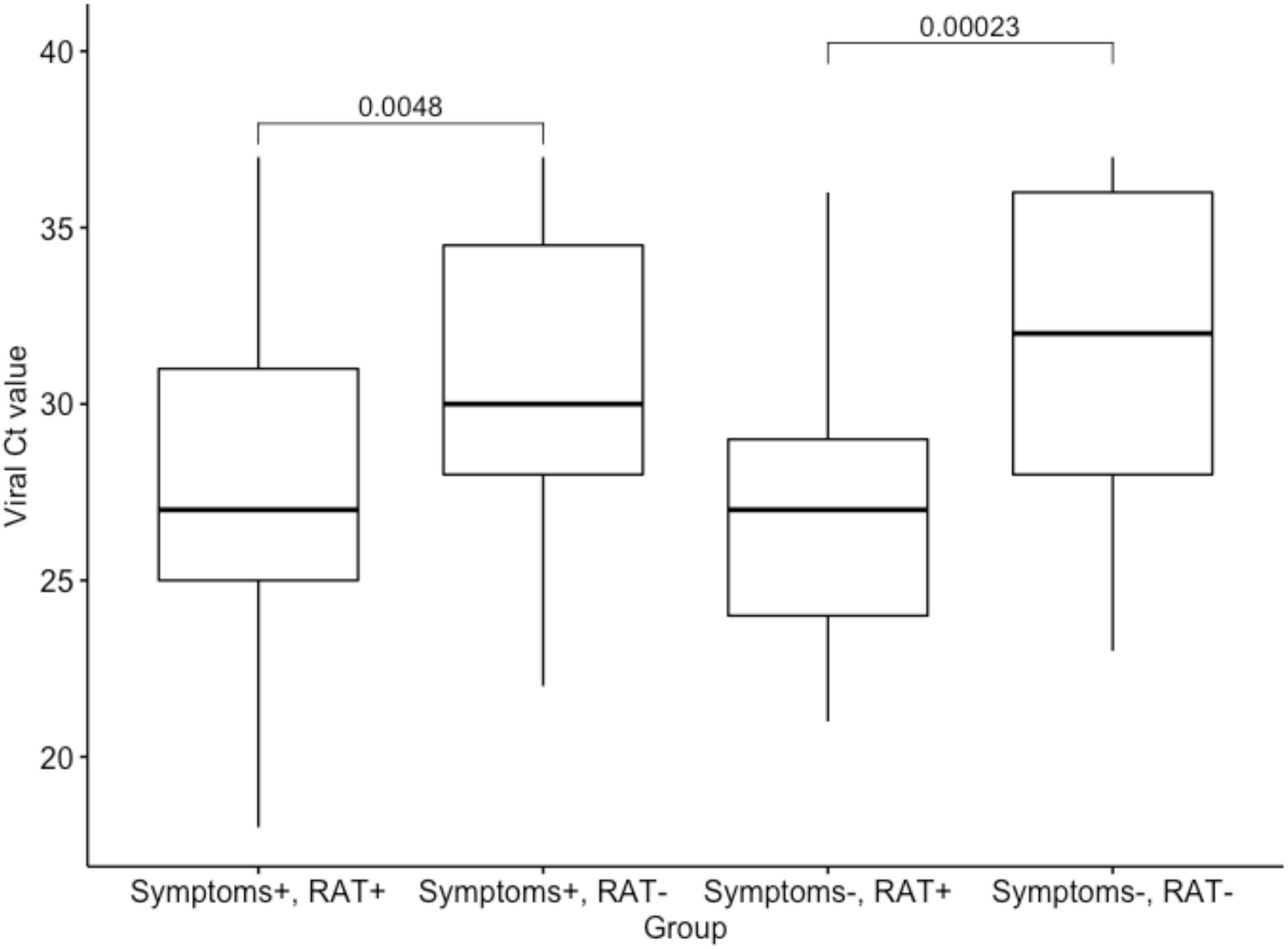
Difference in viral cycle threshold (Ct) value between participants with positive and negative rapid antigen tests among RT-PCR-positive participants with and without self-reported symptoms (n=144). Analysis for statistical difference was performed by Wilcoxon test. Abbreviations: Symptoms+ = individuals with self-reported symptoms, Symptoms- = individuals without self-reported symptoms, RAT+ = positive rapid antigen test, RAT- = negative rapid antigen test

## Discussion

This study comprises a non-selected population with a 4.6% prevalence of SARS-CoV-2 infection. In agreement with recommendation from the Center of Disease Control and Prevention (CDC) on the use of antigen testing, the sensitivity of 69.7% indicates that RATs should not replace RT-PCR in diagnosis and surveillance of SARS-CoV-2 infection^1^. However, the high predictive values, especially in individuals with symptoms, and the fast test result implying faster tracing of infected individuals, supports that RATs can have a significant role in COVID-19 screening. That individuals with false negative results of the RAT had significantly higher Ct value corresponding to a lower viral load, indicate that individuals with false negative RATs are less infectious in general.

The comparison of test results from oropharyngeal and nasopharyngeal swabs suggests a limitation. However, both methods are in accordance with CDC recommendations^2^. Even though RT-PCR is considered the gold standard for detection of SARS-CoV-2 infection, it is not flawless, and the choice of RT-PCR as reference and criteria for positive results have implications^3,4^.

In agreement with WHO’s recommendation of testing for SARS-CoV-2 as intensively as possible, the STANDARD Q COVID-19 Ag test and other RATs with similar accuracy seem to be a good supplement to RT-PCR testing^5^.

## Data Availability

Data are available upon request to the corresponding author, however requires permission from the Danish Authorities which can be applied for.

## Acknowledgements

We thank Copenhagen Medical A/S for delivering the rapid antigen tests and providing test personnel for performing the tests. We thank Andreas Lyman and the personnel at Testcenter Taastrup for their essential assistance in collecting data. Last, we thank Martin Magelund Rasmussen, Anne-Marie Vangsted, and Nikolai Søren Kirkby for critical discussion and help with logistics.

## Author contributions

Ms Jakobsen, Mr. Jensen and Dr. von Buchwald had full access to all the data in the study and take responsibility for the integrity of the data and the accuracy of the data analysis.

Concept and design: Jakobsen, Jensen, Todsen, Klokker, von Buchwald.

Acquisition, analysis, or interpretation of data: All authors.

Drafting of the manuscript: Jakobsen, Jensen.

Critical revision of the manuscript for important intellectual content: All authors.

Statistical analysis: Jakobsen, Jensen.

Administrative, technical, or material support: Jakobsen, Jensen, Todsen, Martel, von Buchwald. Supervision: von Buchwald.

## Conflicts of interest

No conflicts of interest.

## Funding

No funding was received for this project.

## Supplementary

### Methods

ClinicalTrials.gov Identifier: NCT04689399

#### RT-PCR

Detection of SARS-CoV-2 was performed by single target RT-PCR at TestCenter Danmark, Statens Serum Institut. Oropharyngeal swabs were collected by the personnel at Testcenter Taastrup and eluted in PBS and RNA was extracted using RNAdvance Blood (Beckman). One-step RT-PCR to detect SARS-CoV-2 was performed using Luna Universal Probe One-step RT-qPCR kit (New England Biolab). The following primers and probe binding to the E-gene were used:

E_Sarbeco_F (ACAGGTACGTTAATAGTTAATAGCGT),

E_Sarbeco_R (ATATTGCAGCAGTACGCACACA),

E_Sarbeco_P1 (FAM-ACACTAGCCATCCTTACTGCGCTTCG-BHQ1.)

Samples with 10 ≤Ct ≤ 38 were considered positive.

#### Rapid antigen test (RAT)

The WHO EUL-approved STANDARD Q COVID-19 Ag test produced by SD BIOSENSOR was performed by personnel from the private company Copenhagen Medical A/S and conducted according to SD BIOSENSOR’s instructions immediately after the oropharyngeal swab for RT-PCR testing. Participants with non-conclusive RAT tests had a new test performed before leaving the test center until a conclusive result was obtained.

#### Questionnaire

Upon agreeing to participate in the study the participants mobile phone number was registered and link to an online questionnaire was sent by SMS. The questionnaire was developed in REDCap and the participants’ answers were collected here as well. The questions included if the participant:

1. had previously tested positive for COVID-19
2. had been in close contact with a known infected individuals
3. had symptoms of COVID-19

If yes was answered to the third question, the participants were asked which symptoms and for how long he or she had symptoms prior to the test.

#### Ethics

The study followed the Helsinki II Declaration, and all participants gave informed consent. Approval for conducting the study was obtained from the Regional Committee on Health Research Ethics (case nr. 20083631) and the Danish Data Protection Agency (P-2020-1222).

## Notes

### Competing Interest Statement

The authors have declared no competing interest.

### Clinical Trial

ClinicalTrials.gov Identifier: NCT04689399

